# Multimodal Deep Learning Reveals the Modular Genetic Architecture of Cardiovascular Aging

**DOI:** 10.64898/2026.04.22.26351478

**Authors:** Ryan B. Choi, Philip M. Croon, Sudheesha Perera, Lovedeep S. Dhingra, Bruno Batinica, Evangelos K. Oikonomou, Rohan Khera

## Abstract

Age is the dominant risk factor for cardiovascular disease, yet individuals of the same chronological age can differ markedly in the organs and biological pathways through which cardiovascular vulnerability emerges. We used deep learning to estimate biological age from four cardiovascular data streams in more than 100,000 UK Biobank participants - 12-lead electrocardiograms, cardiac magnetic resonance imaging, carotid ultrasound, and retinal fundus photographs - representing electrical, structural, macrovascular, and microvascular domains. The resulting biological age gaps showed limited overlap across modalities in participants with complete phenotyping, and phenome-wide analyses linked each axis to distinct disease profiles. Genome-wide association analyses identified 38 independent loci, with largely modality-specific signals involving electrophysiologic, myocardial structural, vascular regulatory, and ocular/microvascular pathways. Cross-trait LD score regression and polygenic risk scores further supported partial genetic separation of the four axes, with modality-specific associations for atrial fibrillation, heart failure, peripheral arterial disease, hypertension, diabetes, and diabetic retinopathy in UK Biobank and broadly concordant patterns in All of Us. Integration with myocardial single-cell transcriptomic data nominated distinct cellular contexts for these genetic signals. These findings suggest that AI-derived cardiovascular age is not a single biomarker of systemic senescence, but a family of related phenotypes that decompose cardiovascular aging into measurable electrical, myocardial, macrovascular, and microvascular modules.

## Main

Chronological age, the passage of time since birth, is the strongest risk factor for cardiovascular disease.^1^ However, age is distinct from aging, which is a dynamic, highly individualized process of physiological change. Consequently, individuals of the same chronological age often differ markedly in cardiovascular health, reflecting substantial heterogeneity in biological senescence, the progressive cellular and structural degeneration of tissues over time.^1,2^ Biological age has therefore emerged as a complementary construct that aims to capture the cumulative effects of genetics, environmental exposures, and risk factors on tissue and organ function more directly than chronological age alone.^3–7^ The difference between biological and chronological age, often referred to as the biological age gap (BAG), has been associated with adverse outcomes including heart failure, stroke, and mortality, suggesting that accelerated cardiovascular aging represents a clinically meaningful phenotype rather than a passive correlate of time.^8–12^

Recent advances in deep learning (DL) have enabled the estimation of biological age directly from cardiovascular data, generating quantitative biomarkers of aging from modalities such as electrocardiograms (ECG), cardiac magnetic resonance imaging (CMR), carotid ultrasound, and retinal imaging.^9–14^ These approaches are particularly valuable because they can detect latent patterns of physiological decline not readily captured by conventional measurements or human interpretation. As a result, AI-derived age offers a scalable way to quantify cardiovascular age across organ systems and functional domains.

Most prior work has either developed a single modality-specific age clock or combined multiple measurements into a global biological-age index. This has left a central question unresolved: whether an apparently older cardiovascular system reflects one shared aging program, or several partially independent axes that affect the conduction system, myocardium, large vessels, and microvasculature differently. This distinction is important because a composite age score may be clinically useful while still obscuring the biological pathway driving risk in a given individual.

Here, we used AI-derived age gaps from ECG, CMR, carotid ultrasound, and retinal fundus imaging to test whether cardiovascular aging is genetically and clinically modular. We asked three linked questions: whether accelerated aging tends to occur in the same individuals across modalities; whether each modality has distinct genomic architecture and cell-type enrichment; and whether genetic liability to each aging axis maps to different cardiometabolic disease profiles.

## Results

### Study overview

We analyzed multimodal data from 101,621 UK Biobank participants (mean age 63 ± 8.1 years; 54.4% female). There were 77,133 individuals with ECGs, 72,879 with CMR, 72,270 with carotid ultrasound, and 88,084 with retinal imaging. The cohorts for ECG, CMR, and carotid imaging exhibited similar cardiovascular profiles, with mean age of 66 years and prevalence for hypertension (22.3% to 22.9%), type 2 diabetes (4.5% to 4.7%), coronary artery disease (9.5% to 10.0%), and hypercholesterolemia (11.0% to 11.5%) across cohorts. Notably, the fundus imaging cohort was younger (mean age, 58.5 years) with higher burden of cardiovascular comorbidities, most notably hypertension (31.1%) and type 2 diabetes (8.3%) (**Table 1**).

**Table 1.** Table of baseline characteristics.

|  |  | **ECG** | **CMR** | **Carotid** | **Fundus** |
| --- | --- | --- | --- | --- | --- |
| **N** |  | 77133 | 72879 | 72270 | 88084 |
| **Chronological Age** |  | 66.4 ± 7.7 | 66.6 ± 7.7 | 66.1 ± 7.7 | 58.5 ± 8.5 |
| **Biological Age** |  | 65.5 ± 8.2 | 66.4 ± 7.8 | 65.0 ± 7.9 | 59.5 ± 7.8 |
| **Sex (Female)** |  | 40083 (52.0%) | 37842 (51.9%) | 37420 (51.8%) | 47194 (53.6%) |
| **Race** | **White** | 74535 (96.6%) | 70479 (96.7%) | 69952 (96.8%) | 81419 (92.4%) |
|  | **Black** | 597 (0.8%) | 542 (0.7%) | 516 (0.7%) | 2185 (2.5%) |
|  | **Asian** | 935 (1.2%) | 865 (1.2%) | 848 (1.2%) | 2258 (2.6%) |
|  | **Other/unknown** | 829(1.1%) | 760(1.0%) | 729(1.0%) | 1691 (1.9%) |
| **BMI (kg/m<sup>2</sup>)** |  | 26.6 ± 4.3 | 26.6 ± 4.2 | 26.6 ± 4.2 | 27.2 ± 4.7 |
| **Hypertension** |  | 17531 (22.7%) | 16218 (22.3%) | 16561 (22.9%) | 27394 (31.1%) |
| **T2D** |  | 3584 (4.6%) | 3262 (4.5%) | 3399 (4.7%) | 7344 (8.3%) |
| **CAD** |  | 7451 (9.7%) | 6927 (9.5%) | 7195 (10.0%) | 12038 (13.7%) |
| **Hypercholesterolemia** |  | 8780 (11.4%) | 7989 (11.0%) | 8306 (11.5%) | 15232 (17.3%) |

Figure 1 shows the analytic framework. The four AI-age models generated modality-specific BAG phenotypes, which were then analyzed through GWAS in unrelated participants of European genetic ancestry. We evaluated modularity at four levels: phenotypic overlap of accelerated aging, cross-trait genetic correlation, disease associations of modality-specific PRSs in 336,825 UK Biobank participants and 168,878 All of Us participants, and single-cell transcriptomic enrichment.

**Figure 1.**
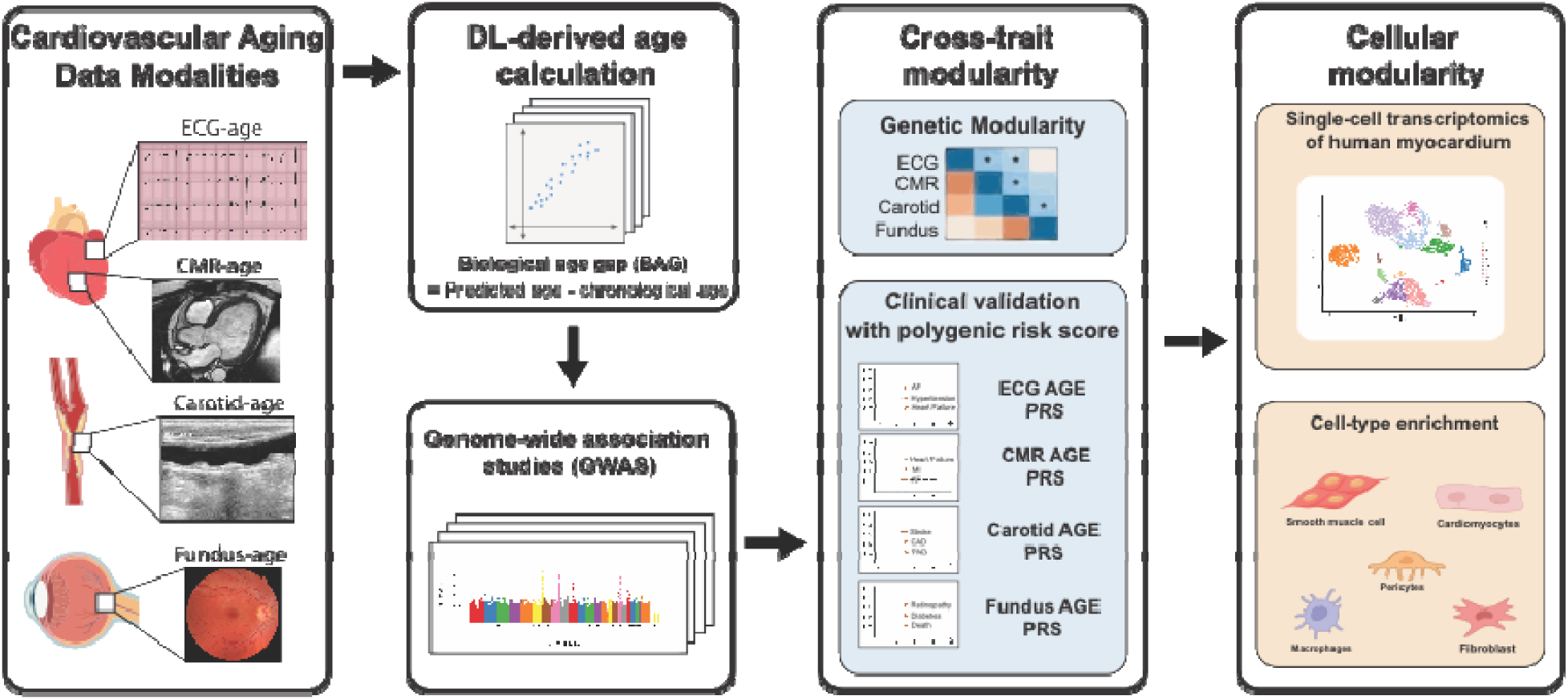
Analytical pipeline for dissecting the modular genetic architecture of cardiovascular aging. Deep learning models calculated biological age gaps across four cardiovascular modalities (ECG, CMR, carotid ultrasound, and retinal imaging) to conduct genome-wide association studies (GWAS). Cross-trait modularity was assessed by mapping genetic correlations and validating clinical disease trajectories via polygenic risk scores (PRS). Finally, genomic risk loci were integrated with myocardial single-cell transcriptomics to identify the specific cell-type enrichments driving each aging module. *ECG, electrocardiogram; CMR, cardiac magnetic resonance; DL, deep learning; PRS, polygenic risk score.*

### Deep learning captures age prediction across cardiovascular modalities

The AI-age models, each trained to minimize the mean absolute error (MAE) against chronological age using modality-specific deep learning architectures, estimated age with performance that varied across modalities (Figure 2A). The CMR model had the lowest MAE (4.28 years; R2=0.75), followed by fundus imaging (4.71 years; R2=0.74), carotid ultrasound (5.71 years; R2=0.58), and ECG (5.91 years; R2=0.58). To characterize multi-domain aging burden, we defined accelerated aging within each modality as a BAG exceeding that modality’s own MAE, a harmonized threshold that accounts for the differing predictive accuracy across data types. Among participants with complete data across all four modalities (n=15,343), only 153 (1.0%) met the accelerated-aging threshold in all four modalities, whereas 2,225 (14.5%) did so in two or more modalities (Figure 2B). Accelerated aging therefore most often appeared in a modality-specific pattern rather than as pan-cardiovascular acceleration.

**Figure 2.**
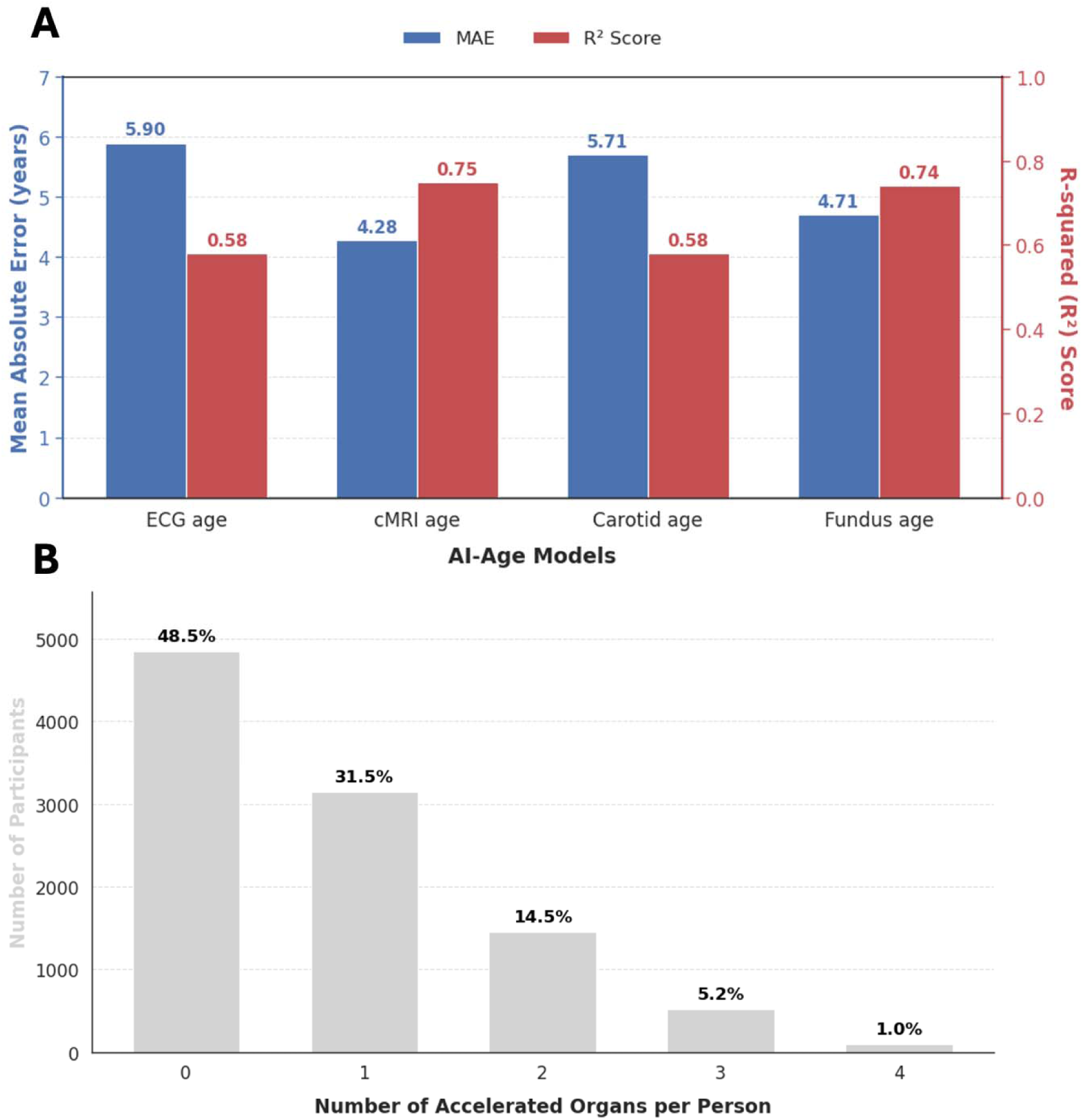
AI-derived biological age model performance and multi-domain aging burden. (A) Mean absolute error (MAE, years; blue) and R2 (red) for each AI-age model across ECG, CMR, carotid, and fundus modalities in UK Biobank. (B) Distribution of the number of concurrently accelerated aging domains per participant among individuals with complete data across all four modalities; accelerated aging was defined as BAG greater than the modality-specific MAE. ECG, electrocardiogram; CMR, cardiac magnetic resonance.

To systematically characterize the clinical correlates of each BAG phenotype, we performed phenome-wide association studies (PheWAS) linking accelerated aging status in each modality to ICD-10 coded diagnoses from linked NHS health records. This identified 108, 179, 72, and 189 FDR-significant disease associations for ECG, CMR, carotid, and fundus accelerated aging, respectively. ECG acceleration was distinctly enriched for cardiac arrhythmias and conduction disorders (I44: OR=3.79; I48: OR=2.27), CMR for cardiometabolic comorbidities (I50: OR=1.86; E11: OR=1.84), carotid for vascular risk factors and respiratory disease (F10: OR=2.12; J44: OR=1.94), and fundus for ocular phenotypes and diabetic complications (H36: OR=3.57; E11: OR=1.67) as shown in the **supplementary figures**. These FDR-significant hits demonstrated minimal overlap across the four modalities, with each aging metric displaying a distinct distribution of disease chapters. The shared variance in significant phenome-wide associations between any two modalities was low, with pairwise overlap ranging from 22.3% (carotid-fundus) to 42.1% (ECG-CMR), and only 33 codes shared across all four modalities.

### Genome-wide discovery identifies modality-specific loci across cardiovascular aging axes

To identify genetic determinants of the continuous modality-specific age gaps, we performed GWAS for each BAG phenotype (Figure 3A). Across the four modalities, we identified 38 independent genome-wide significant loci: 15 for ECG-age, 7 for CMR-age, 7 for carotid-age, and 9 for fundus-age.

**Figure 3.**
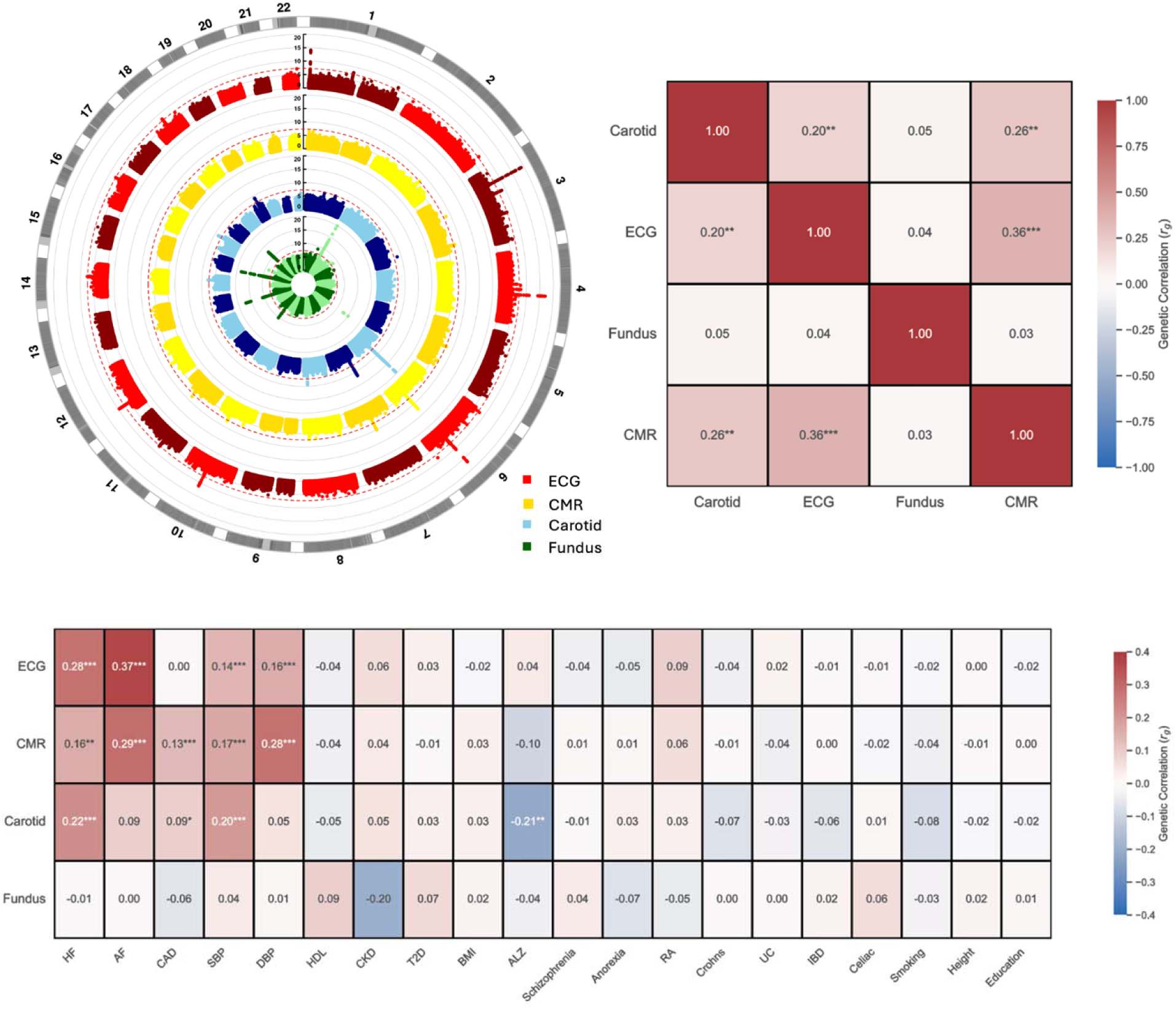
Genetic architecture and cross-trait correlations of **cardiac** aging modalities. (A) Circular Manhattan plot displaying GWAS results for all four aging modalities (ECG age, red; cMRI age, yellow; carotid age, blue; fundus age, green), with chromosomal position on the outer ring and –log_₁₀_(p) on the radial axis. Dashed red line denotes genome-wide significance (P=5×10^−8^). (B) Pairwise genetic correlation matrix (rg) between the four aging modalities estimated by bivariate LD score regression. (C) Cross-trait genetic correlations between each aging modality and 20 established clinical GWAS traits. Color intensity reflects rg; asterisks denote statistical significance (*p<0.05, **p<0.01, ***p<0.001). HF, heart failure; AF, atrial fibrillation; CAD, coronary artery disease; SBP, systolic blood pressure; DBP, diastolic blood pressure; HDL, high-density lipoprotein; CKD, chronic kidney disease; T2D, type 2 diabetes; BMI, body mass index; ALZ, Alzheimer disease; RA, rheumatoid arthritis; UC, ulcerative colitis; IBD, inflammatory bowel disease.

ECG-age loci converged on electrophysiological stability and excitation-contraction coupling. The strongest signal mapped to the *SCN5A/SCN10A* locus (rs6801957, p=2.31×10⁻²³), which governs action potential depolarization via the primary cardiac sodium channel. Additional signals underscore the importance of sarcoplasmic reticulum calcium cycling in electrical aging, including *CAMK2D* (p=1.01×10⁻¹□), *CASQ2* (p=1.28×10⁻□), and *PLN* (p=2.47×10⁻□), alongside *PRDM16* (p=6.93×10⁻¹□), a transcription factor implicated in metabolic and structural cardiomyopathies. eQTL mapping confirmed cardiac tissue-specific regulatory effects.

CMR-age loci were highly enriched for the maintenance of sarcomeric integrity, myocardial elasticity, and mechanotransduction. These loci were anchored by *PLN* (rs72967533, p=3.50×10⁻¹³) and *TTN* (p=4.10×10⁻□), the primary molecular determinant of passive myocardial stiffness. Signals at *ELN* (p=8.18×10⁻¹¹) and *ANKRD1* (p=2.23×10⁻□) further implicate extracellular matrix remodeling and mechanical stress-response programs in gross structural aging.

Carotid-age loci exhibited a distinct intergenic-dominant architecture (enrichment=1.33, p=5.34×10⁻²¹) with depletion from exonic regions, consistent with distal transcriptional regulation of vascular gene programs. These signals mapped to loci governing endothelial function and macrovascular remodeling, notably including *VASH1*, an endothelium-derived angiogenesis inhibitor, and the *BCAR1/CFDP1* locus, which is implicated in endothelial mechanotransduction and arterial stiffness, alongside *ELOVL4/BCKDHB* and *CEBPA*.

Fundus-age loci showed protein-coding enrichment (exonic enrichment=2.67, p=9.73×10^−5^) and implicated ocular and microvascular biology. Loci mapped near GJA3/GJB2, connexin genes involved in intercellular communication, and KCNQ1, a potassium-channel locus with vascular and metabolic associations. Other signals, including TSPAN10, OCA2/HERC2, IRF4, and SH3YL1/ACP1 (p=1.02×10^−59^), likely capture a mixture of retinal, pigmentation, and microvascular phenotypes; these loci therefore require careful interpretation in relation to image-derived age. Additional annotation for each GWAS is provided in the Supplementary Materials.

### Modular genetic architectures across cardiovascular aging axes

To quantify the degree of shared genetic architecture among the four aging phenotypes, we performed bivariate LD score regression across modalities (Figure 3B). CMR-age showed moderate genetic overlap with ECG-age (rg=0.36) and carotid-age (rg=0.26), whereas ECG-age and carotid-age showed weaker correlation (rg=0.20). Fundus-age showed no significant genetic overlap with the other modalities, supporting partial modularity rather than a single shared genetic aging factor.

We calculated cross-trait genetic correlations (*r_g_*) between the four AI-aging phenotypes and 20 clinical trait GWAS summary statistics using LDSC (Figure 3B). ECG-age demonstrated significant genetic correlation with atrial fibrillation (*r_g_* = 0.37, P < 0.001), heart failure (*r_g_* = 0.28, P < 0.001), systolic blood pressure (*r_g_*= 0.14, P < 0.001), and diastolic blood pressure (*r_g_* = 0.16, P < 0.001). Similarly, CMR-age was genetically correlated with atrial fibrillation (*r_g_* = 0.29, P < 0.001), heart failure (*r_g_* = 0.16, P < 0.01), coronary artery disease (*r_g_* = 0.13, P < 0.001), systolic blood pressure (*r_g_* = 0.17, P < 0.001), and diastolic blood pressure (*r_g_* = 0.28, P < 0.001).

Carotid-age shared genetic variance with heart failure (*r_g_* = 0.22, P < 0.001) and systolic blood pressure (*r_g_* = 0.20, P < 0.001). In contrast, fundus-age demonstrated no statistically significant genetic correlations across the 20 examined phenotypes, including cardiovascular, metabolic, psychiatric, autoimmune, and anthropometric traits.

### Modality-specific polygenic risk scores associate with distinct cardiometabolic outcomes

To test whether modality-specific genetic liability had clinically interpretable correlates, we examined associations between each PRS and nine cardiometabolic outcomes in UK Biobank and All of Us (Figure 4C). In UK Biobank, the ECG-age PRS was most strongly associated with atrial fibrillation (OR per SD=1.09), with additional associations for heart failure, stroke, and hypertension. The CMR-age PRS was associated with all evaluated outcomes, including atrial fibrillation (OR per SD=1.10), heart failure (OR per SD=1.05), stroke (OR per SD=1.05), and MACE (OR per SD=1.05). The carotid-age PRS was associated with peripheral arterial disease (OR per SD=1.05) and hypertension (OR per SD=1.05), whereas the fundus-age PRS had its strongest associations with diabetic retinopathy (OR per SD=1.09) and diabetes (OR per SD=1.05).

**Figure 4.**
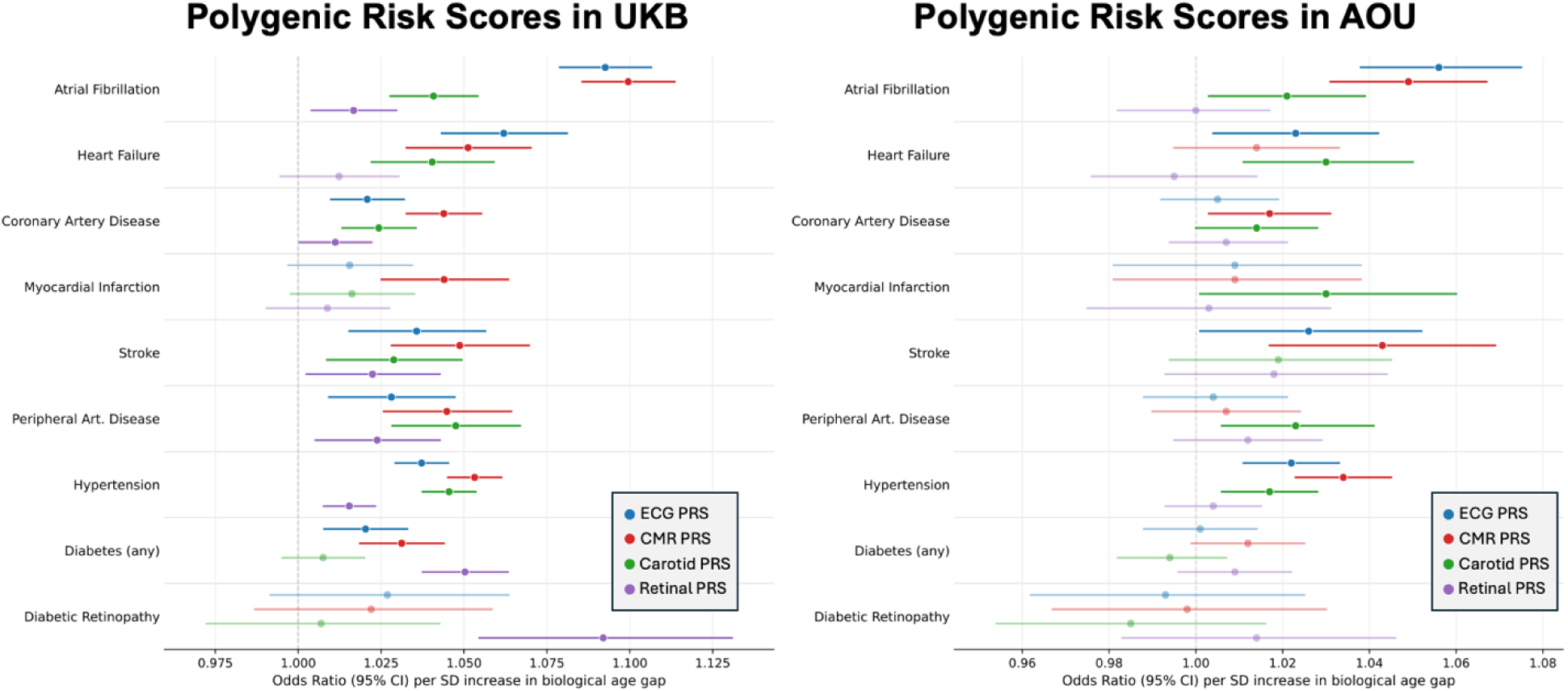
Polygenic risk score associations with cardiovascular and metabolic outcomes. Forest plot of odds ratios (ORs, 95% CIs) per one-standard-deviation increase in PRS for each of the four cardiovascular aging modalities across nine clinical outcomes in UK Biobank (n=336,825) and All of Us (n=168,878). Models were adjusted for age at recruitment, sex, and the first ten genetic principal components. Faded markers denote associations that did not reach statistical significance (p>=0.05). Dashed vertical line indicates OR=1.0.

In All of Us, effect directions were concordant for 83% of trait-outcome pairs. The strongest UK Biobank associations--including ECG-age PRS with atrial fibrillation, heart failure, and stroke, and carotid-age PRS with peripheral arterial disease and hypertension--remained statistically significant. Several associations for lower-prevalence outcomes, including fundus-age PRS with diabetic retinopathy and diabetes, were directionally consistent but did not reach significance, emphasizing that these PRS results should be interpreted as genetic corroboration of modality-specific biology rather than as clinically deployable risk scores.

### Single-cell transcriptomic enrichment nominates distinct myocardial cell populations for each cardiovascular aging axis

We applied scDRS to 166,636 cells from a curated human myocardium atlas using MAGMA-derived gene scores from each aging GWAS (Figure 5). Each modality showed a different myocardial cell-type enrichment profile. ECG-age enrichment localized to cardiomyocytes (z=3.94, P=0.001) and smooth muscle cells (z=3.30, P=0.003). CMR-age showed enrichment in cardiac neurons (z=3.01, P=0.006) and cardiomyocytes (z=2.38, P=0.010). Carotid-age demonstrated enrichment in endocardial cells (z=3.35, P=0.002), B cells (z=2.99, P=0.004), and endothelial cells (z=2.53, P=0.008). Fundus-age showed broader myocardial enrichment across fibroblasts, smooth muscle cells, cardiac neurons, NK/T cells, and adipocytes. These results nominate possible cellular contexts for genetic signals, while recognizing that retina- and vascular-specific atlases will be needed to confirm tissue-matched mechanisms for fundus and carotid aging.

**Figure 5.**
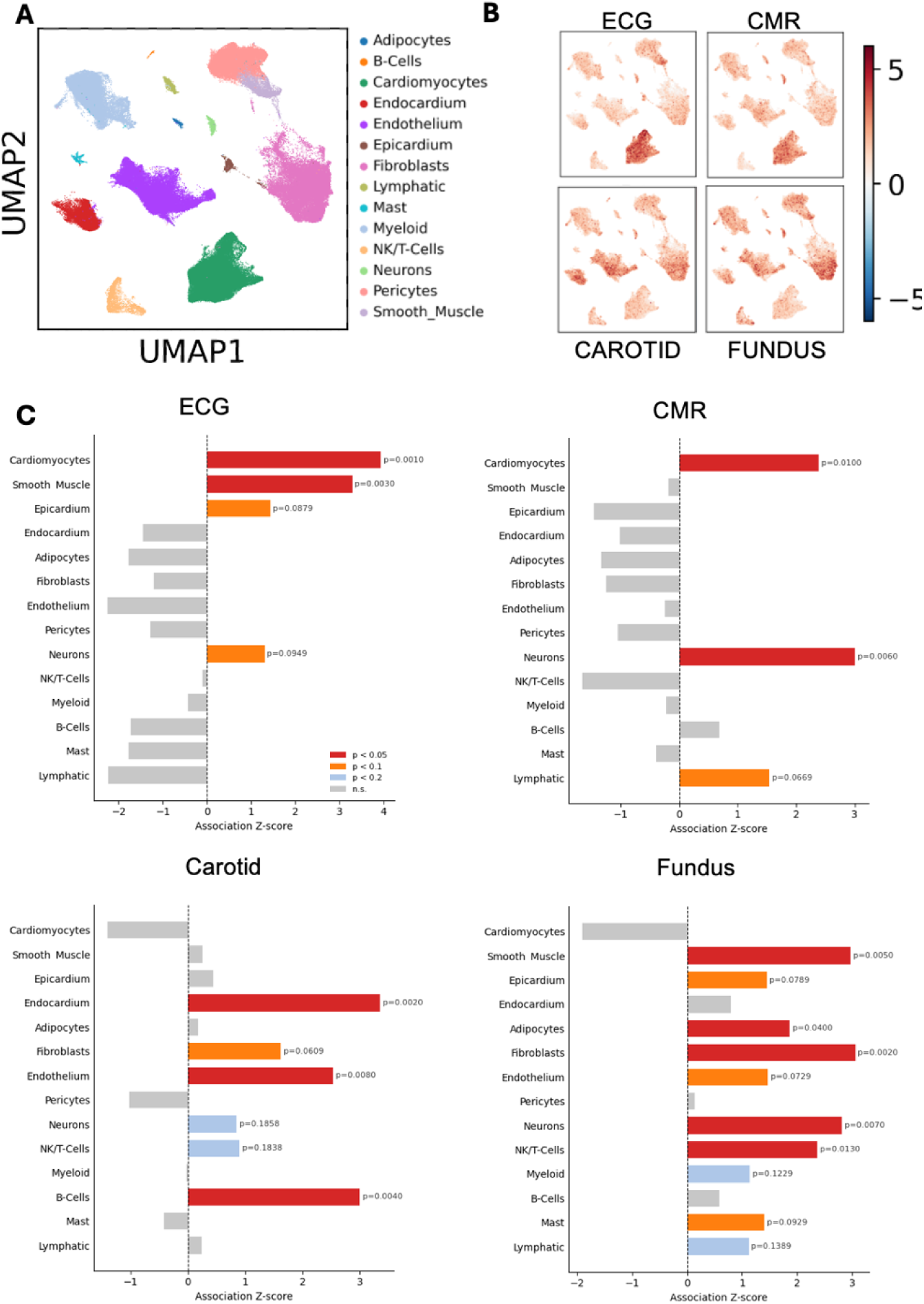
scDRS cell-type association analysis linking cardiovascular aging GWAS signals to human myocardial single-cell transcriptomes. (A) UMAP embedding of the cardiac single-cell transcriptomic atlas colored by annotated cell type, comprising 14 major populations. (B) UMAP feature plots of per-cell scDRS disease scores for ECG-age, CMR-age, carotid-age, and fundus-age. (C) Horizontal bar plots showing scDRS cell-type association z-scores for each aging trait; bars are colored by significance threshold and annotated with exact p values for nominally associated cell types. Cell-type order is consistent across panels to facilitate comparison.

## Discussion

In this study, AI-derived age from ECG, CMR, carotid ultrasound, and retinal fundus imaging decomposed cardiovascular aging into partly distinct physiological and genetic axes.

Accelerated aging most often occurred in one or a small number of modalities rather than across all four. GWAS identified largely modality-specific loci, LDSC showed only partial genetic overlap, PRS analyses linked each axis to different cardiometabolic phenotypes, and single-cell analyses nominated different myocardial cell contexts. Together, these findings argue against a single, synchronized cardiovascular aging clock and instead support a modular model in which electrical, myocardial structural, macrovascular, and microvascular aging can progress at different rates within the same individual.

These findings extend molecular aging-clock literature by focusing on tissue-level readouts of cardiovascular structure and function. Epigenetic, proteomic, metabolomic, and transcriptomic clocks can capture systemic molecular aging, whereas imaging- and signal-derived clocks may reflect accumulated remodeling in specific cardiovascular territories.^15–18^ The present multimodal decomposition suggests that these measures are not interchangeable: ECG-age preferentially indexes electrical and excitation-contraction biology, CMR-age indexes myocardial structure and mechanics, carotid-age indexes macrovascular remodeling, and fundus-age indexes ocular and microvascular features.

Framing cardiovascular age as modular is important because composite age indices can conceal heterogeneity.^19,20^ A single elevated cardiovascular age score might arise from conduction-system remodeling, ventricular structural change, large-vessel disease, or microvascular pathology. In contrast, modality-specific age gaps provide a way to map the pathway through which aging-related vulnerability is expressed. This framework may also explain why aging clocks trained on different data sources sometimes show only modest agreement despite similar performance for chronological age prediction.

The strongest contribution of the current study is biological stratification: the same chronological age can conceal different domains of cardiovascular vulnerability, and genetic analyses suggest that these domains have different upstream determinants. With further validation, modality-specific age gaps could help refine risk phenotyping, prioritize preventive evaluation, and provide intermediate phenotypes for genetic discovery. However, clinical use would require evidence that these measures improve decisions beyond chronological age and established risk models, are robust across acquisition protocols and populations, and lead to actionable changes in care.

Several limitations should be considered. First, these imaging- and signal-based clocks are not ready for clinical deployment; the models require external validation across imaging vendors, acquisition protocols, health systems, and ancestrally diverse populations. Second, UK Biobank imaging participants are a selected volunteer population, and the fundus cohort differed in age and comorbidity burden from the other modality cohorts, which may influence cross-modality comparisons. Third, BAG phenotypes are dynamic and shaped by environment, disease, and measurement context; germline analyses capture only the inherited component and cannot establish causality. Fourth, fundus loci that map to pigmentation-related genes raise the possibility that some retinal age signal reflects image or ocular phenotype rather than vascular aging alone. Fifth, myocardial single-cell enrichment provides biological plausibility for cardiac modalities but is not tissue-matched for carotid or retinal phenotypes. Finally, any multimodal AI framework must show incremental value beyond chronological age and conventional risk models before it can be justified in practice.

In our findings, multimodal deep learning shows that cardiovascular aging can be resolved into distinct but partially overlapping electrical, myocardial structural, macrovascular, and microvascular axes. The modality-specific genetic architectures, disease associations, and cellular enrichments observed here provide a framework for studying cardiovascular aging as a set of modular biological processes rather than a single chronological trajectory.

## Methods

All analyses were conducted under the approved UK Biobank project ID #71033, with multimodal data exported to a secure Yale University-managed server under a formal data transfer agreement. Because this study involved the secondary analysis of pre-existing, de-identified data, it was deemed exempt from full review by the Institutional Review Board at Yale University.

### Overview

Deep learning models were trained to estimate chronological age from four cardiovascular modalities: 12-lead ECGs, CMR imaging, carotid ultrasound, and retinal fundus photography. For each participant, the biological age gap (BAG) was calculated as the difference between model-predicted age and chronological age, yielding four modality-specific quantitative phenotypes. These phenotypes were then used for GWAS, cross-trait genetic correlation analyses, PRS construction and validation, and integration with single-cell transcriptomic data.

### Data sources and study population

We analyzed data from UK Biobank participants, a national cohort study in the UK with extensive health and biometric data through linkage to the National Health Service (NHS) electronic health records. All individuals have available genotyping data. For this study, we included all participants with at least one cardiovascular phenotyping modality, spanning 12-lead ECG, CMR, carotid ultrasound, or retinal fundus photography.^21^ All participants provided written consent, and those who subsequently withdrew consent were excluded from the analysis.

For external validation of the PRS, we additionally analyzed data from the All of Us (AOU) Research Program (Controlled Tier dataset v8),^22^ a US-based longitudinal cohort with linked electronic health record and short-read whole-genome sequencing data. The validation cohort was restricted to participants of European genetic ancestry to match the ancestry composition of the PRS derivation cohorts, and associations between each PRS and prevalent cardiovascular and metabolic diagnoses were assessed using diagnosis codes from linked EHR data. As with the UK Biobank cohort, all included participants had provided consent for research use of their electronic health record and genomic data.

### Preprocessing of included data

We analyzed four complementary cardiovascular phenotyping modalities: 12-lead ECG, CMR long-axis cine imaging, carotid ultrasound, and retinal fundus photography. ECG signals, a recording of cardiac conduction, were sampled at 500 Hz over 10 seconds across 12 leads and standardized by median centering. For CMR, a representation of cardiac structure and function, we implemented a centering and masking algorithm to isolate cardiac motion, synchronizing 2-, 3-, and 4-chamber long axis views into a 3D spatiotemporal video. Carotid duplex assessments, a view of an important macrovascular territory, were distilled into maximal-variance frames to capture vascular hypertrophy and calcification. Fundoscopic images of the retina were standardized for luminance and contrast enhancement to highlight microvascular structures. Full technical parameters and preprocessing pipelines are detailed in the **supplementary methods.**

### Development of the AI-Aging Models

We used a unified training protocol across all modalities to ensure that differences in model behavior reflected the underlying data rather than variation in modeling strategy. For each data modality, we established a training cohort of approximately 5,000 randomly selected individuals, stratified into equal age bins to maintain a uniform chronological age distribution. We trained all models to minimize the mean absolute error (MAE) using the AdamW optimizer (learning rate = 1 × 10^−3^). Detailed methodology protocols are provided in the **supplementary methods.**

### Phenotyping accelerated aging across multiple modalities

We defined accelerated aging within each modality as a BAG greater than that modality-specific MAE. This threshold was used as a harmonized, modality-specific cut point for descriptive overlap analyses rather than as a clinical diagnostic threshold. To compare burden across systems, we restricted this analysis to participants with complete data for all four modalities and calculated an accelerated-aging count ranging from 0 to 4. We then performed a Phenome-Wide Association Study (PheWAS) to determine whether the number and type of accelerated-aging domains were associated with differential disease burden (**Supplementary Methods**).

### Genome-wide association studies

We conducted GWASs across all four cardiovascular data modalities using REGENIE.^23^ The primary quantitative phenotype for association testing was modality-specific BAG. To minimize genetic confounding, stringent quality control restricted the analysis to unrelated individuals of European ancestry, explicitly excluding participants with genetic sex discordance or sex chromosome aneuploidy.

### LDSC-cross-trait heritability

Genetic architecture overlap was quantified through bivariate LD score regression (LDSC)^24^ to estimate cross-trait heritability. We estimated genetic correlation (*r_g_*) between our four AI-derived aging phenotypes and a curated set of publicly available GWAS summary statistics. These comparators included a variety of cardiovascular diseases, chronic conditions, and biometric traits.

### Annotations of variants

Genome-wide significant loci were functionally annotated and mapped to genes using functional mapping and annotation (FUMA).^25^ Independent genomic risk loci were defined by lead SNPs (5 × 10^-8)^ clustered within a 250kb window (r^2^ < 0.6). We prioritized putative causal genes using a multi-modal mapping strategy: (1) Positional mapping where genes were located within 10kb of the lead SNP; (2) eQTL Mapping in which genes where the lead SNP is a significant *cis*-eQTL (FDR < 0.05) in relevant cardiovascular tissues (e.g., Artery Aorta, Heart Left Ventricle) from GTEx v8; (3) Chromatin Interaction where genes physically linked to the locus via 3D chromatin loops (Hi-C) in cardiac tissue.

### Polygenic risk scores

PRSs for the four aging modalities were constructed with PRSice-2 (v2.3.5)^26^ to test whether inherited liability to each AI-derived age gap was associated with downstream cardiometabolic phenotypes. PRS were developed from the original GWAS developmental UKB cohorts for each respective cardiovascular modality. Target analyses were performed in UK Biobank participants who were withheld from the original AI age-model training, as well as in an external, independent AOU cohort. Both target cohorts were restricted to individuals of matching European ancestry to minimize population stratification. Clumping used a 250-kb window, an LD threshold of r2 < 0.1, and a clumping p-value of 1. Allele harmonization, including matching effect alleles, correcting strand mismatches, and excluding ambiguous palindromic variants, was performed automatically by PRSice-2 between the base and target datasets. Variants were included at a prespecified p < 0.05 threshold, yielding approximately 300,000 independent variants per modality. This fixed threshold was chosen to avoid optimization on the validation outcome; all PRSs were standardized to z-scores before association testing with nine cardiovascular and metabolic outcomes.

### Single cell enrichment

Modality-specific genetic signal was integrated with single-cell RNA sequencing (scRNA-seq) data from the human heart.^27^ We constructed a healthy reference atlas by re-processing raw transcriptomic data using Seurat v5.^28^ We applied the Single-Cell Disease Relevance Score (scDRS) framework^29^ to map polygenic signatures of cardiovascular aging to cellular populations. MAGMA^30^ was used to prioritize gene-level associations and scDRS was used to project these polygenic signals onto myocardial single-cell atlas, computing a normalized disease relevance score for each individual cell based on its transcriptomic profile. We then computed Pearson correlation coefficients between individual gene expression matrices and the continuous cell-level scDRS scores, prioritizing genes that strongly associate with modality-specific aging risk.

### Statistical analysis

Deep learning models were trained to minimize MAE relative to chronological age, and model fit was evaluated using Pearson correlation coefficients and R2. The modality-specific MAE was used only to define accelerated aging for overlap and PheWAS analyses. GWAS for continuous modality-specific BAG used linear regression adjusted for age, sex, genotyping array, and the first ten genetic PCs, with genome-wide significance defined as P < 5 × 10^−8^. LDSC and scDRS analyses used FDR correction. PRS-outcome associations were assessed with multivariable logistic regression adjusted for baseline age, sex, and the first ten PCs. All tests were two-sided.

## Data availability

Individual-level data for the UK Biobank cohort are not publicly available due to participant privacy and consent restrictions. Qualified researchers can request access through the UK Biobank Access Management System following approval of a research application. Individual-level data from the All of Us Research Program are similarly not publicly available; qualified researchers may apply for access to the Controlled Tier dataset through the All of Us Researcher Workbench following institutional registration and completion of required training. Modality-specific GWAS summary statistics generated during this study will be deposited in the NHGRI-EBI GWAS Catalog or another appropriate public repository upon publication.

## Code Availability

Custom analytical code and pipelines used for deep learning model training, GWAS processing, PRS construction, and single-cell enrichment analyses will be made publicly available upon publication. Code will be deposited in a public repository at that time.

## Acknowledgements

R.B.C. was supported by a fellowship from the Sarnoff Cardiovascular Research Foundation.

## Author Contributions

R.K. conceived and supervised the study. R.B.C., P.M.C., S.P., L.S.D., B.B., and E.K.O. performed the analyses and interpreted the data. R.B.C. and R.K. drafted the manuscript. All authors critically revised the manuscript for important intellectual content and approved the final version.

